# A rapid review of patient and family perspectives on inappropriateness of intensive care treatments at the end of life

**DOI:** 10.1101/19007138

**Authors:** Magnolia Cardona, Shantiban Shanmugam, Ebony T Lewis, Alex Psirides, Matthew Anstey, Ken Hillman

**Affiliations:** Institute for Evidence-Based Healthcare, Faculty of Health Sciences and Medicine, Bond University., 14 University Drive, Robina Campus, QLD 4229, Australia; Gold Coast Hospital and Health Service., 1 Hospital Blvd, Southport, QLD, 4215, Australia; Concord Hospital, Sydney., Hospital Rd, Concord NSW, 2139, Australia; School of Public Health and Community Medicine, The University of New South Wales., Samuels Avenue, Kensington NSW 2033, Australia; Intensive Care Unit, Wellington Regional Hospital., Riddiford St, Newtown, Wellington 6021, New Zealand; Intensive Care Medicine, Sir Charles Gairdner Hospital., Hospital Ave, Nedlands WA 6009, Australia; Intensive Care Unit, Liverpool Hospital., Elizabeth &, Goulburn St, Liverpool NSW 2170, Australia; South Western Sydney Clinical School, The University of New South Wales., Goulburn St, Liverpool NSW 2170, Australia

**Keywords:** professional-patient relations, intensive care, end-of-life, review

## Abstract

**Aim:** To understand patient/family perspective of inappropriate intensive care unit (ICU) admissions and treatment.

**Methods:** Rapid literature review of English language articles published between 2001 and 2017 in Medline or PsycInfo.

**Results:** Thirteen articles covering 6,194 elderly patients or surrogate decision-makers from four countries were eligible. Perceived inappropriateness of ICU treatments was mainly expressed as dissatisfaction with clinicians’ as surrogate decision-makers, inconsistency with patient/family values, family distrust of physician’s predictions on poor prognosis, and inadequate communication on over-aggressive treatment causing suffering. Consultation on opinion before ICU admission varied from 1% to 53.6%, and treatment goals from 1.4 to 31.7%. Satisfaction with the decision-making process in ICU was higher for those who had certain level of control and involvement in the process.

**Conclusions:** The patient/family perspective on inappropriateness of ICU treatments involves preferences, values and social constructs beyond medical criteria. Earlier consultation with families before ICU admission, and patient education on outcomes of life-sustaining therapies may help reconcile these provider-patient disagreements.

**Take-home message:** The patient/family perspective on *inappropriateness* of ICU at the end of life often differs from the clinician’s opinion due to the non-medical frame of mind. To improve satisfaction with communication on treatment goals, consultation on patient values and inclusion of social constructs in addition to clinical prediction are a good start to reconcile differences between physician and health service users’ viewpoint.

## Background

Elderly patients at the end of life are high consumers of hospital and intensive care unit (ICU) services, yet their outcomes remain poor, many die during admission and others die within months of ICU discharge. [1, [2] Moreover, of those that survive an ICU stay, their long term quality of life and physical health are often severely compromised after hospital discharge. [3] High quality end-of-life care in ICU has been defined by families as encompassing timely and compassionate communication, shared decision-making incorporating patient treatment goals and values, avoidance of prolongation of dying, and preservation of comfort and dignity. [4] Despite this, with increasingly available technology to prolong life -the medicalisation of death- [5] has resulted in many of these ICU admissions deemed *inappropriate* from a medical perspective [6] and in some cases unwanted by the patients themselves, who would largely prefer less aggressive treatments at the end of life. [7]

Clinicians may generally have some understanding of what constitutes *inappropriate* care at the end of life. [8] Treatments are considered medically *inappropriate* due to many factors: the intensity of resource use deemed more substantial than warranted, [9] patients being too ill to benefit from ICU management, [10] unnecessary treatments when there is no hope of surviving the ICU stay, [11] the intervention is expected to have a negligible impact on recovery of independence, [8] or the treatment having no bearing on the health outcome or quality of life of the person. [12] Yet doctors still administer aggressive treatments to patients at the end of life, [13] even though recognition of dying has occurred. [14] This may create confusion among patients’ and family’s understanding of the expected trajectory. It is then not surprising that no consensus on the patient or family perception of what constitute *appropriate* admission to ICU or *appropriate* treatment in an ICU are available.

A first step in decision-making about continuing or suspending aggressive care implies the delivery of negative prognostic information to families, [15] a practice that more often than not involves spending more time speaking to families and less time listening to their concerns. [16, [17] The central theme of these conversations is clarifying the treatments goals. That is, deciding whether the aim of treatment is curative, palliative, or terminal depending on the expected response and clinical outcomes and giving the patient or family opportunity to decline treatments if they so wish. [18] Patients or families are often asked to participate in decisions to withdraw or withhold treatment after a patient has been admitted to ICU [19, [20] but conflict over the end-of-life care in ICU and strategies to resolve this conflict are still a concern. [21]

To our knowledge there has been no accepted definition of what intensivists agree is appropriate. Limited research also appears to be available on the health service user perspective. [8, [22, [23] Hence we conducted this review in an attempt to understand the concept and illustrate the perception of *inappropriateness* of ICU treatments from the patient and family perspective when there was no longer a true prospect of benefit for the patients but a possibility of treatment causing more harm than good.

## Specific objectives

This review aims to address the following questions:

1. To what extent are older patients and their families involved in decisions about *appropriateness* of intensive care admission or treatments?
2. How do patients/families define *inappropriate* intensive care admission or treatments?

## Methods

We searched for English language articles published from January 2001 to November 2017 in two databases: Medline and PsycInfo. For details of the methods search, study types, eligibility criteria and data extraction see Supplement 1.

## Results

Thirteen publications produced over the past 16 years and covering 6,194 patients or surrogate decision-makers addressed at least one of our research questions (Table 1). The eight quantitative, four qualitative and one legal report from five countries (9 from USA, 2 from Canada, 1 Australia and 1 from France-Hungary) mainly targeted surrogate decision-makers (10/13) but half (7/13) also involved elderly patients themselves. We found only two studies directly offering the patient or family definitions of or a perspective on *inappropriateness* of treatment at the end of life, [12, [24] one study sought to determine if elderly patients were consulted before admission to ICU, [25]and one study specifically addressing *inappropriateness* of care in ICU. [26] All other studies mostly investigated satisfaction with end-of-life care or discordance between healthcare providers and families about the need for or benefits of certain ICU procedures or treatments.

**Table 1.**
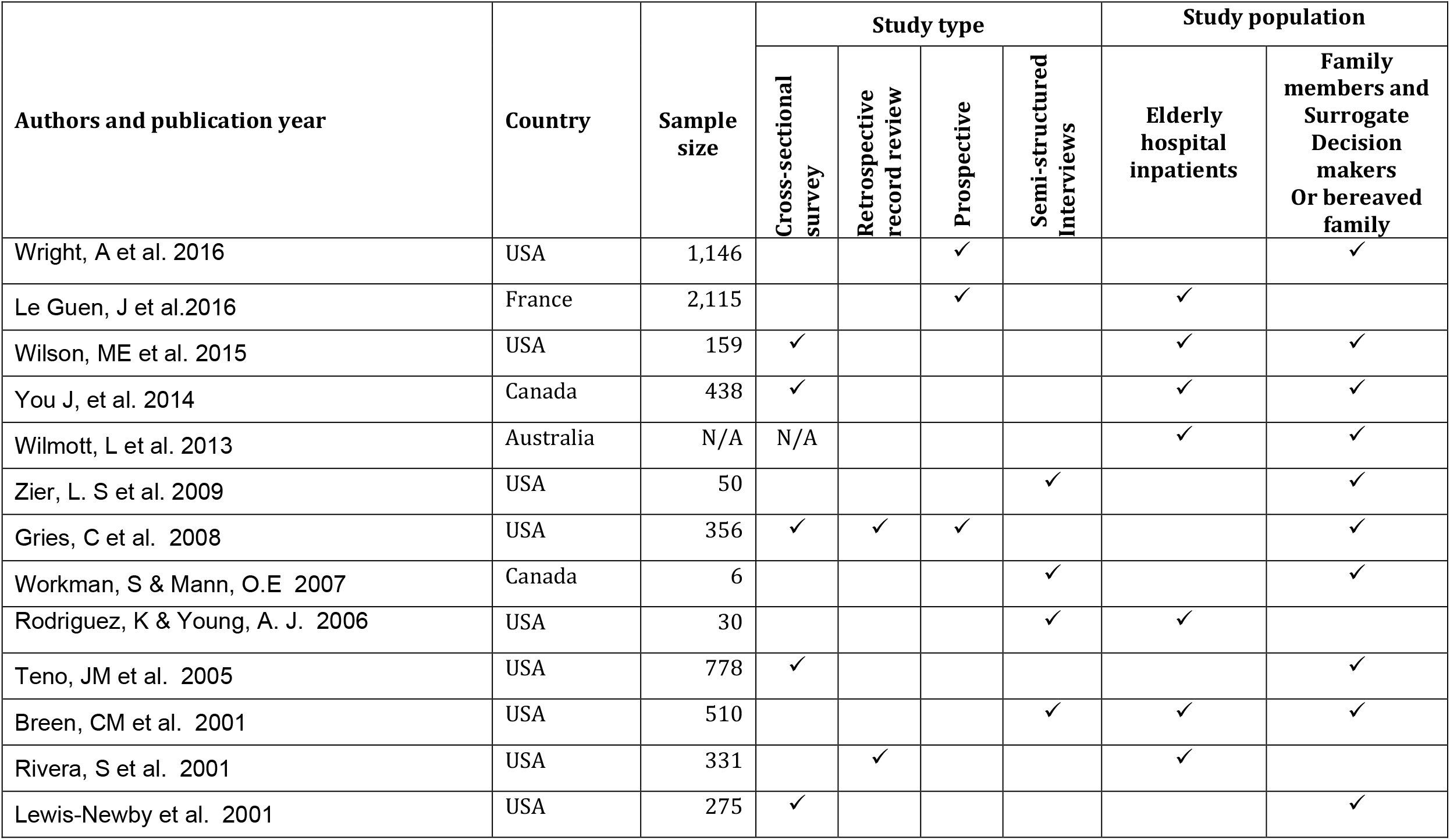
Relevant studies, methods and study populations – 2000 to 2016.

| Authors and publication year | Country | Sample size | Study type |  |  |  | Study population |  |
| --- | --- | --- | --- | --- | --- | --- | --- | --- |
|  |  |  | Cross-sectional survey | Retrospective record review | Prospective | Semi-structured Interviews | Elderly hospital inpatients | Family members and Surrogate Decision makers Or bereaved family |
| Wright, A et al. 2016 | USA | 1,146 |  |  | ✓ |  |  | ✓ |
| Le Guen, J et al.2016 | France | 2,115 |  |  | ✓ |  | ✓ |  |
| Wilson, ME et al. 2015 | USA | 159 | ✓ |  |  |  | ✓ | ✓ |
| You J, et al. 2014 | Canada | 438 | ✓ |  |  |  | ✓ | ✓ |
| Wilmott, L et al. 2013 | Australia | N/A | N/A |  |  |  | ✓ | ✓ |
| Zier, L. S et al. 2009 | USA | 50 |  |  |  | ✓ |  | ✓ |
| Gries, C et al. 2008 | USA | 356 | ✓ | ✓ | ✓ |  |  | ✓ |
| Workman, S & Mann, O.E 2007 | Canada | 6 |  |  |  | ✓ |  | ✓ |
| Rodriguez, K & Young, A. J. 2006 | USA | 30 |  |  |  | ✓ | ✓ |  |
| Teno, JM et al. 2005 | USA | 778 | ✓ |  |  |  |  | ✓ |
| Breen, CM et al. 2001 | USA | 510 |  |  |  | ✓ | ✓ | ✓ |
| Rivera, S et al. 2001 | USA | 331 |  | ✓ |  |  | ✓ |  |
| Lewis-Newby et al. 2001 | USA | 275 | ✓ |  |  |  |  | ✓ |

In the absence of a published definition in most studies, and due to the reported significant association between inappropriateness and dissatisfaction with medical treatments in ICU (p=0.0003) [26] we accepted dissatisfaction measurements as a proxy definition of *appropriateness* from the health service user perspective. Search results and exclusions based on title and abstract are presented in the PRISMA diagram (Supplement 1).

### Patient/family definitions

The terms *inappropriate* or *futile* were generally not used in the reviewed studies but a recent survey of ICU patients and their surrogates in the US and Hungary reported that “too much treatment” was perceived inappropriate mainly due to misalignment with either patient or family wishes, or because it caused unacceptable suffering, or was too costly. [26] A small fraction (5.4%) of respondents deemed some ICU interventions *inappropriate* by this definition.

In a small US veteran’s clinic study, elderly outpatients defined *futility* as those treatments administered when the patient “has nothing to look forward to”, or “is a vegetable”, or the treatments are “a waste of time and money”. [12] Factors predisposing participants to categorise end-of-life interventions as *futile* were a low likelihood of treatment success, a limited expected effect on patient’s longevity and quality of life, and an anticipated emotional and financial cost to the family. [12]

A smaller qualitative study also reported that families felt more in control if patients were admitted to ICU where care, although traumatic, is more personalised and perceived to be of higher quality. By contrast, palliative care was perceived as abandonment of patients or providing interventions to hasten their death. [27] From this health service user perspective, survival was more important than palliation and thus aggressive care in the ICU was paradoxically associated with higher levels of surrogate satisfaction. [27] On the other hand, bereaved family members in a large retrospective study of Medicare beneficiaries, were less likely to consider admission to an ICU within 30 days of death for patients with advanced stage cancer as excellent quality of care. [28] This view was associated with the place of death not being consistent with the patient’s wishes.

### Extent of patient/family consultation

Consulting patients about their perceptions, values, fears, risks and preferences was not commonplace (Table 3). Lack of consultation with patients or lack of control over decision on treatment discontinuation by families (5/13 studies) or inadequate communication (4/13 studies) were reported to cause dissatisfaction with ICU management (Table 2). The extent of satisfaction with family involvement varied depending on the completeness of information given to them and on the coverage of the priority issues in the communication.

**Table 2.** Domains covered by the definitions of perceived inappropriateness of/dissatisfaction with ICU management.

| Authors,<br>publication year and<br>study type | Inconsistent<br>with patient/<br>family values | Not<br>accepting<br>prognosis/<br>conflict with<br>prediction | Lack of<br>consultation with<br>patient/ lack of<br>control by family | Inadequate<br>Communication<sup>§</sup> | Too<br>aggressive<br>/causing<br>suffering | Cost | Lack of<br>understanding of<br>limitations of life-sustaining<br>therapies | Lengthy<br>ICU stay |
| --- | --- | --- | --- | --- | --- | --- | --- | --- |
| **Quantitative studies** |  |  |  |  |  |  |  |  |
| Le Guen 2016 |  |  | ✓ |  |  |  |  |  |
| Wright 2016 | ✓ |  |  |  | ✓ |  |  | ✓ |
| Wilson 2015 | ✓ |  |  |  | ✓ | ✓ |  |  |
| You 2014 |  |  |  | ✓ |  |  |  |  |
| Lewis-Newby 2011 |  |  | ✓ | ✓ |  |  |  |  |
| Gries 2008 | ✓ | ✓ | ✓ |  |  |  |  |  |
| Teno 2005 | ✓ |  | ✓ | ✓ | ✓ |  |  |  |
| Breen 2001 |  | ✓ |  |  |  |  |  |  |
| Rivera 2001 | ✓ | ✓ |  |  |  |  |  |  |
| **Qualitative studies** |  |  |  |  |  |  |  |  |
| Zier 2009 | ✓ | ✓ |  |  |  |  |  |  |
| Willmott 2013 | ✓ |  |  |  |  |  |  |  |
| Workman 2007 |  |  | ✓ | ✓ |  |  | ✓ |  |
| Rodriguez 2006 | ✓ |  |  |  | ✓ | ✓ |  |  |

<sup>§</sup> includes treatment options, consequences, preferences, treatment goals or values

**Table 3.**
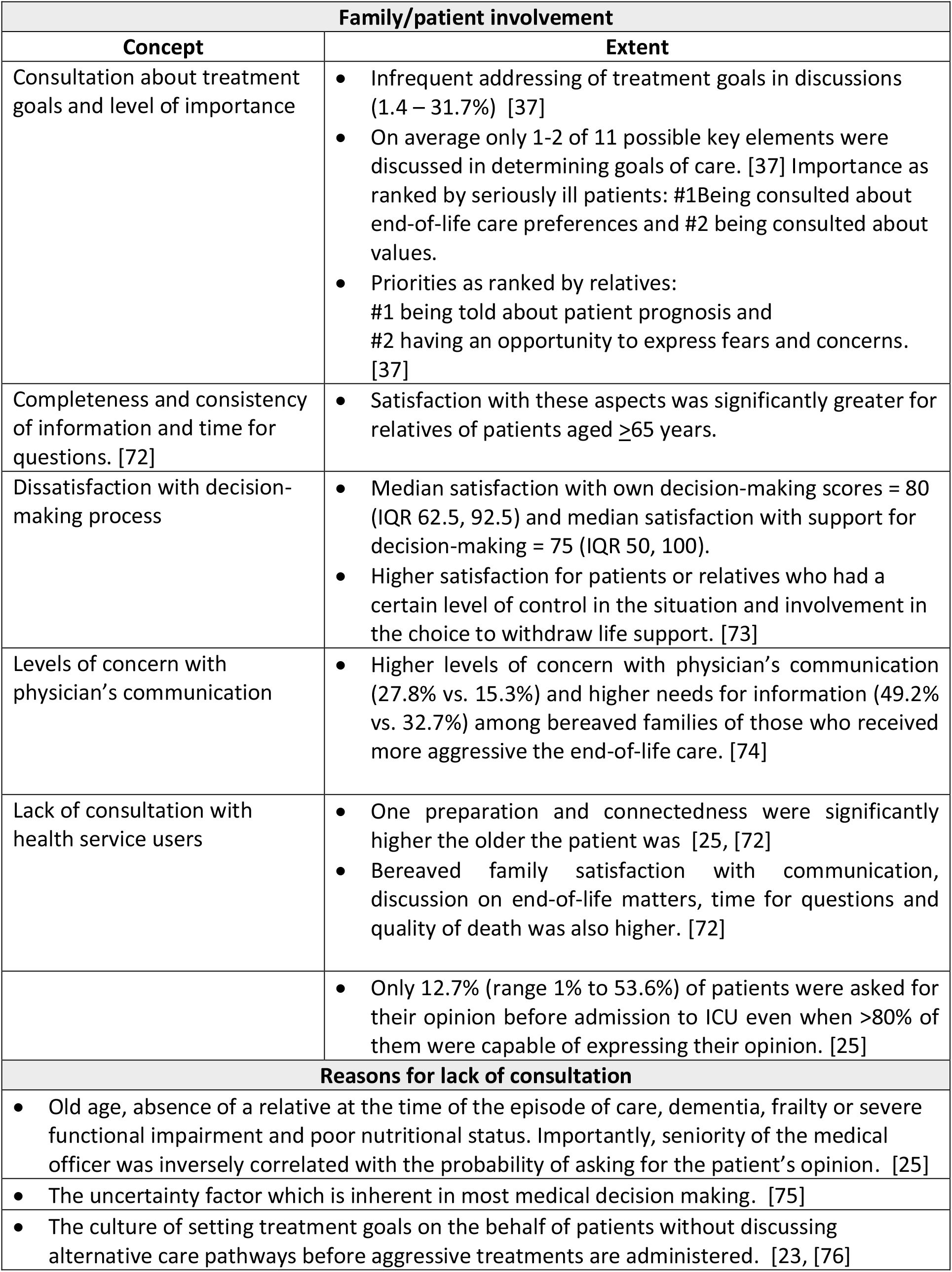
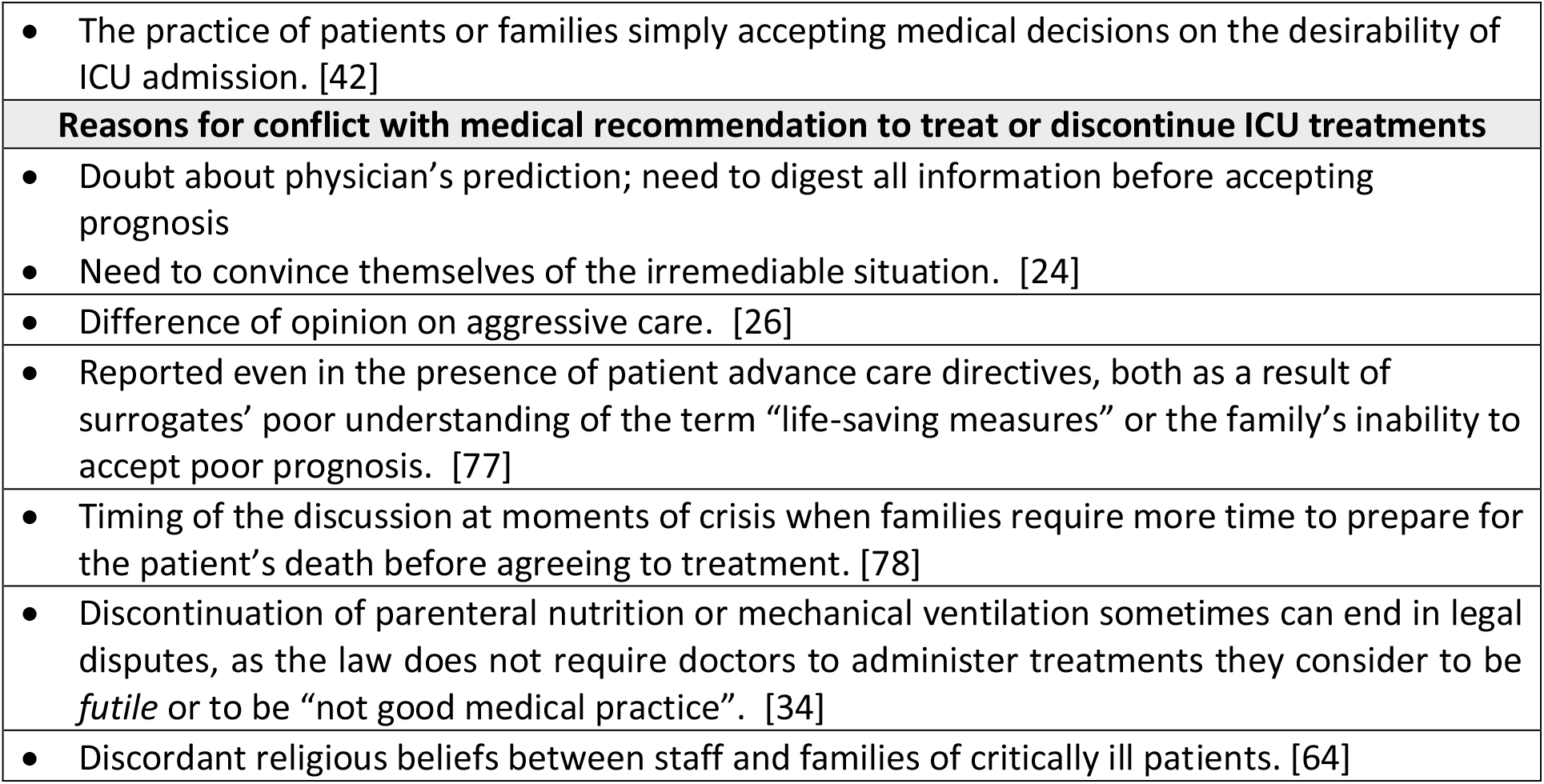
Extent of family involvement and factors contributing to communication failures between healthcare providers and patients/families.

While prognostic issues before admission to ICU should be discussed with health service users, [29, [30] the role of initiating the end-of-life discussion or consultation on patient preferences for transfer to IC may vary across health systems from the emergency physician on admission in some, to the bioethics expert or the palliative care team during hospitalisation in others. [31, [32, [33] These grey areas can sometimes have serious emotional and legal implications [34] as decisions are usually carried out from a medical perspective. Discussion on preferences or rationale for ICU admission can be affected by the patient’s mental competence to communicate preference; [25] the predicament of doctors to witness the grief of relatives while balancing prognostic estimates and decision making; [35] and the family preparedness to accept the inevitable without attempting further care. [36] Other diverse factors contributing to communication failure regarding prognosis are shown in Table 3.

### Health care service user-provider discordance

Even after discussions on treatment goals take place, there is a risk that insufficient communication can lead to conflict if prognosis, values, care preferences, and fears or concerns are not adequately addressed. [37] Family disagreement with health professional advice, hereby assumed as disagreement with the medical definition of inappropriateness, was reported in 5 publications. The most commonly cited domain of patient dissatisfaction with (inappropriateness of) ICU treatment was inconsistency with patient or family values (8/13 publications, Table 2). Refusal to accept prognosis (4/13) and aggressive care causing suffering (4/13) were also reported as contributors of dissatisfaction with ICU management. The least frequently reported reasons for dissatisfaction with ICU treatments were high cost (2/13), presumable due to differential in funding arrangements across health systems, where in some cases costs to the individual may not be relevant. Family disagreements due to prolonged length of ICU stay or lack of understanding of the aim of limitations of life-sustaining therapies were also rarely reported (1/13 each).

Overall in our review the main reasons for dissatisfaction with ICU care were inconsistency of medical decisions and consumer values, poor understanding of the aims of life-sustaining therapy and lack of acceptance of poor prognosis (Tables 2 and 3).

## Discussion

The patient/family perspective on *inappropriateness* of ICU interventions at the end of life has had limited attention in the medical and nursing literature, but the few studies found confirm that there is a difference in the interpretation of inappropriateness between health service users and healthcare providers. Actual definitions and reports of the extent of patient/family perceived inappropriateness are scarce but based on the few articles found it is clear that *inappropriateness* encompasses factors such as personal and religious beliefs, alignment with patient preferences and values, quality of life post-intervention, treatment costs, and emotional impact.

The terms *satisfaction* with healthcare and *quality of death* are more often investigated, and usually measured in terms of scores or scales.

Reported dissatisfaction with selected ICU treatments originated predominantly from the discordance between medical decisions and consumer values rather than from lack of consultation. Patient/caregivers were found to describe ‘inappropriate’ as not being consistent with their wishes, whereas clinicians generally describe it from a knowledge-base where the delivered intensive procedures exceeded expected benefit [38] does not match the patient’s poor prognosis or the treatment has no impact on quality of remaining life. [39, [40, [41] As end-of-life decisions to withdraw or withhold treatments from patients who cannot communicate for themselves, [42] are usually delegated to surrogates who may be unaware of patients wishes and therefore request futile treatments, [43] the discordance between the medical and family constructs reflects the difference in that knowledge base. This dissonance may be the reason for the unrealistic expectation by families that the purpose of ICU is to *attempt everything* including prolongation of care and death with the advent of technology. *[36]*

It is acknowledged that some places may be conducting Intensive communication with ICU patients and relatives and this has shown to be associated with lasting reductions in ICU length of stay and reduced mortality in critically ill adult medical patients. [44] Yet there is no indication that the practice is widespread. In some countries (including the USA, Australia and New Zealand) doctors are not obliged to seek consent from surrogate decision-makers or patients to cease inappropriate treatments. [45, [46] Instead they have long been considered to be in first line to determine appropriateness or futility [47] as they are by default acting in the best interest of patients. As such families do not have the legal right to insist on specific interventions. [34] In many cases the urgency of the decision required can trample these good intentions if a prior end-of-life discussion has not been held. [25] The dilemma is that not arriving at a joint decision contravenes the principle of patient autonomy, [12] but If the patient is incapacitated, the autonomy cannot be delegated unless a family member who possesses the relevant legal right through a power of Attorney to act in the patient’s best interest for health matters. [46, [48]

The growing demand for ICU admission for the very elderly and a need to respect patient autonomy, [25] are acknowledged, as loss of self-determination is associated with perceived loss of control of the dying process and corresponding loss of dignity. [49] The question emerging is whether there is a point where a balance or satisfactory negotiation can be achieved on the extent of ICU care or decision to admit to ICU without prolonging suffering and death. The ideal situation would be where the decision is neither doctor-driven nor patient/surrogate-driven [45] but a reconciliation of the two perspectives. Patient and family priorities such as number of available staff or ability to interact with family while admitted to hospital may influence desirability of ward admission over ICU admission, reflecting lack of understanding in general populations of the type of care required for the level of illness severity. [50] Further, family involvement in critical decisions in ICU is known to lead to psychological distress, anxiety and depression regardless of whether patients die or survive the ICU admission. [51] Hence, consultations on appropriateness of ICU treatments for terminal patients may be undertaken early to give families time to observe the deterioration and adjust to the inevitable; [52] and detailed information provided with caution to certain subpopulations who are not fully informed of the concepts of severity or the consequences of their involvement in decision-making.

When there are discordant opinions between families and health professionals, decisions to admit the dying elderly for short periods to trial care in ICU in an attempt to address the discordance. [53] This practice is motivated by prognostic uncertainty, the medicalisation of death and professional fear of failure. [54, [55] Clinicians may advocate for these time-limited ICU intensive treatments until prognosis is more certain or until differences of opinion within families are reconciled as families realise the treatment goals need to change from curative to palliative. [53] Paternalism is said to detract from the ethical obligation of involving families in ICU decisions [56] but lack of consultation may reflect the nature of the ICU environment where patients are critically ill and cannot be consulted and their surrogate decision-makers may not be present. [57] Is there an alternative? Consultation before ICU admission, if feasible, could contribute to less dissatisfaction as it is known that the timing of negative prognostic communication is associated with the preparedness of families and the complexity of decision-making [15] While many families accept recommendations for withdrawing or withholding treatment, [42] unreasonable expectation of recovery still contributes to patient or family requests for medically inappropriate treatments. [24] On the other hand, some relatives of patients on long dying trajectories only seek information and reassurance rather than direct input into decisions whilst others prefer to only decide on treatment goals and place of death rather than choose specific treatments. [58] For surrogates wanting an honest prognosis and an involvement in decision-making, communication should go beyond the passing of information [59] and into consultation and support.

## A way forward

Given the domains considered in the reviewed studies, we suggest that the concept of patient/family perceived *inappropriateness* of ICU admission or treatments should cover both the medical and social constructs. That is, evidence of consultation, inclusion of patient values and preferences, resolution of patient-physician discordance and measures of family satisfaction with quality of death. If the main reasons for discontinuing treatments such as poor prognosis, perceived non-beneficial therapies and patient preference, [60] were communicated to families before admission and formalised in an advance care plan, [61] patients and families may be able to better grasp the concept of inappropriateness and better fulfil their role in representing the patient’s best interest.

It is feasible to interview or survey bereaved relatives and even patients in ICU about this topic and more research is required into the trade-offs that the community will be willing to accept. Much education on the difference between withdrawing, withholding, and allowing to die [62] is still needed to improve public understanding of the role and implications of palliative and comfort care and the potential inappropriateness of ICU admission. More importantly, education is also essential to enhance medical and nursing understanding of these differences and prevent conflict within the profession and between professionals and the public. In the face of some certainty of irreversibility, the end-of-life transition in ICU is a coordinated effort and in the medical world orders for limitations of life-sustaining treatment do not necessarily imply abandonment of patient care. [63] However, when it is clear the treatment goals will not be achieved, prolonging life is a violation of ethical principles [64].

Strategies that have been shown to reassure families of the appropriateness of the ICU treatments and generate more satisfaction include: obtaining consent, [65] better step-by-step communication updates as status and indications for treatment changes, sufficient time for information exchanges, consistent information and knowing the role of each service provider. [66] While surviving the ICU to discharge is heralded as a ‘success’ even if from the patient’s perspective, communicating the expected long-term quality of life after ICU discharge may in theory assist families in making informed decisions that match the patient’s personal values and treatment goals. [67] This prognostic information is not usually disclosed (or known) by intensivists and extrapolation from observational datasets may amount to ‘best guess’.

Health service user consultations in this review were short of proposals for strategies to negotiate disagreements between the scientific and the belief-based sections of the community on the time and the type of end-of-life interventions. Other gaps identified by us and others for future research include: the perception by different decision-maker roles within a family; the rationale for discordance with the treating team; investigation of variations in perception of inappropriateness by socioeconomic level and across cultures; and evaluation of the effect of interventions to increase listening by critical care clinicians.

## Limitations

Most of the reviewed studies did not offer a definition *inappropriateness* from the patient/family perspective or analysed the results by population subgroups such as ethnic minorities or various health systems. Satisfaction with involvement in decision-making rather than its extent was generally reported. This may have been due to our searches been limited to two major databases (Medline and PsycInfo). The generalisability of the findings is limited by the few articles found, publications predominantly from English speaking countries, and largely ethnically homogeneous participants. It is possible that we missed some keyword in our online search but we also conducted extensive manual searches of reference lists for eligible articles.

## Implications for practice

It is clear that using medical criteria alone for decision-making on ICU transfers at the end of life carries the ethical dilemma of overlooking patient values and preferences. [68] In routine practice it is feasible to consult families of critically ill patients or bereaved relatives [35] and it would be useful to document patient/family preference for the balance between quality and quantity of life and consult them on how this perception changes with age or progressive disability.

One role of the physician is to deliver prognostic news sensitively, help families accept the imminence of death, involve families in decision-making when the patient is incapacitated, and co-ordinate healthcare providers in the effective application of end-of-life-care extending to limitations of life-sustaining treatment. [69] This could be assisted by recognising and effectively using the informal roles of family members as they emerge during crisis situations. These have been identified as Primary Care Giver, Primary Decision Maker, Family Spokesperson, Out-of-Towner, Patient Wishes Expert, Protector, Vulnerable Member, and Health Care Expert. [70] Consultation with them can minimise conflict and facilitate negotiations on prevention of inappropriate ICU admission or interventions.

The family conference or other approaches to joint decision-making are opportunities for reconciling differences of opinion and for enhancing families understanding of the consequences of *inappropriate* ICU care at the end of life. It is important to understand factors beyond religious objections that make surrogate decision-makers disagree with medical recommendations and insist on interventions when the chances of survival are extremely low or non-existent. Our findings on what matters to patients and families can help design education or communication programs for clinicians and families. For over two decades guidelines have stated that restricting ICU admission for the elderly is unethical if the decision is made on the grounds of age. [25, [71] However, limitations of life support are shown to be more acceptable for them and their families, [72] and probably this debate needs to be reignited to include the prospects of recovery, functional autonomy and quality of life the elderly patient will have following discharge. [25] Drawing the line on *medically* inappropriate ICU care could start with discussions within the various treating specialists about those elderly patients who have multiple factors for poor prognosis to reach consensus on whether the time is right for withdrawing or withholding treatments. Conflict resolution with families can involve compassionate approaches to managing requests for potentially inappropriate treatments, understanding what the family hopes to achieve, and explaining realistic expectations of the benefit of aggressive interventions. [32, [62]

## Conclusions

Perceived *inappropriateness* of ICU treatments for families and patients is multifactorial and it involves social constructs beyond the medical rationale. Health service users appreciate consultation on their values and improved communication for shared decisions about ICU admissions and treatments, but their definition of *inappropriateness* appears to clash with the goal-of-treatment orientation of the medical perspective. Discordance between healthcare provider and health care user perceptions and satisfaction with end of life management in ICU continues to be a consistent finding across studies. Much work lies ahead in clinicians’ understanding the family experience and consulting families before ICU admission to reconcile these differences.

## Data Availability

All data available is presented including supplement

## Author contributions

KH conceived the research question. MC designed the study protocol, conducted independent online searches in parallel, cross-checked eligibility/ exclusions, contributed manual searches, prepared manuscript tables and wrote the first and all subsequent drafts incorporating all co-authors comments. SS conducted the online and manual literature searches and eligibility assessment in parallel, prepared some of the manuscript tables and contributed intellectual input to manuscript versions. EL, AP, MA and KH contributed clinical commentary to interpretation of results, provided references for the discussion and reviewed all versions of the manuscript. All authors approved the final version.

## SUPPLEMENT 1 – DETAILS OF RAPID REVIEW METHODS

### Data sources, eligibility and PICOS criteria

For articles targeting **(P)** older patients (aged 60 years and above) with advanced or terminal illness admitted to ICU and/or their families or caregivers. **(I)** Anticipated articles for review were those investigating any approaches or practices with evidence that the treating team disclosed details of risks and benefits of treatment options and possible unexpected effects and/or consulted family/patient perspective or preference on admission to ICU. They were included in the review if they addressed at least one of our two research questions. **(C)** Comparator (if relevant) were patient groups or settings where no discussion had been held on patient perspective on appropriateness of admission to or treatment in ICU. **(O)** Outcomes of interest included but were not limited to:

- Proportions reporting involvement (opportunity) in decision to admit to ICU
- Proportions involved in/consulted on acceptability of ICU treatments
- Self-reported or proxy-reported appropriateness/desirability of ICU admission
- Family or patient satisfaction or regret with ICU admission or treatment
- Definitions of *inappropriate* from the patient or family viewpoint

**(S)** Study types eligible for this review were all qualitative or quantitative: Delphi studies, focus groups, in-depth interviews, qualitative studies and systematic reviews of qualitative studies; and quantitative studies such as cohort and cross-sectional surveys included only if the aim or outcomes were perceived inappropriateness of ICU admission or treatment. The following were agreed for exclusion: Case studies, duplicate studies, editorial articles unless they were literature reviews on the topic, studies with a focus on clinicians’ perceived inappropriateness, and studies or outcomes where inappropriate was defined as “too little” care.

### Screening and data extraction

Searches were conducted simultaneously by two authors (SS and MC) and inclusion or exclusions for all papers were assessed by one author (SS) and cross-checked by another (MC). Discrepancies were discussed and resolved with the involvement of a third colleague with expertise in literature reviews.

### Search Strategy

Search terms as follows were eligible if included in title or abstract only: “End of life OR Terminal OR Advanced chronic illness OR Dying” AND “Intensive care OR ICU” AND “Patient preference OR Decision making OR Surrogate decision OR Treatment Options OR Famil$” AND “Inappropriate$. OR Futil$ OR Non-beneficial OR Cost-ineffective OR Disproportionate OR Perceived OR Satisfaction OR ICU Outcomes OR Survival OR mortality”.

## SCREENSHOT OF MEDLINE SEARCH

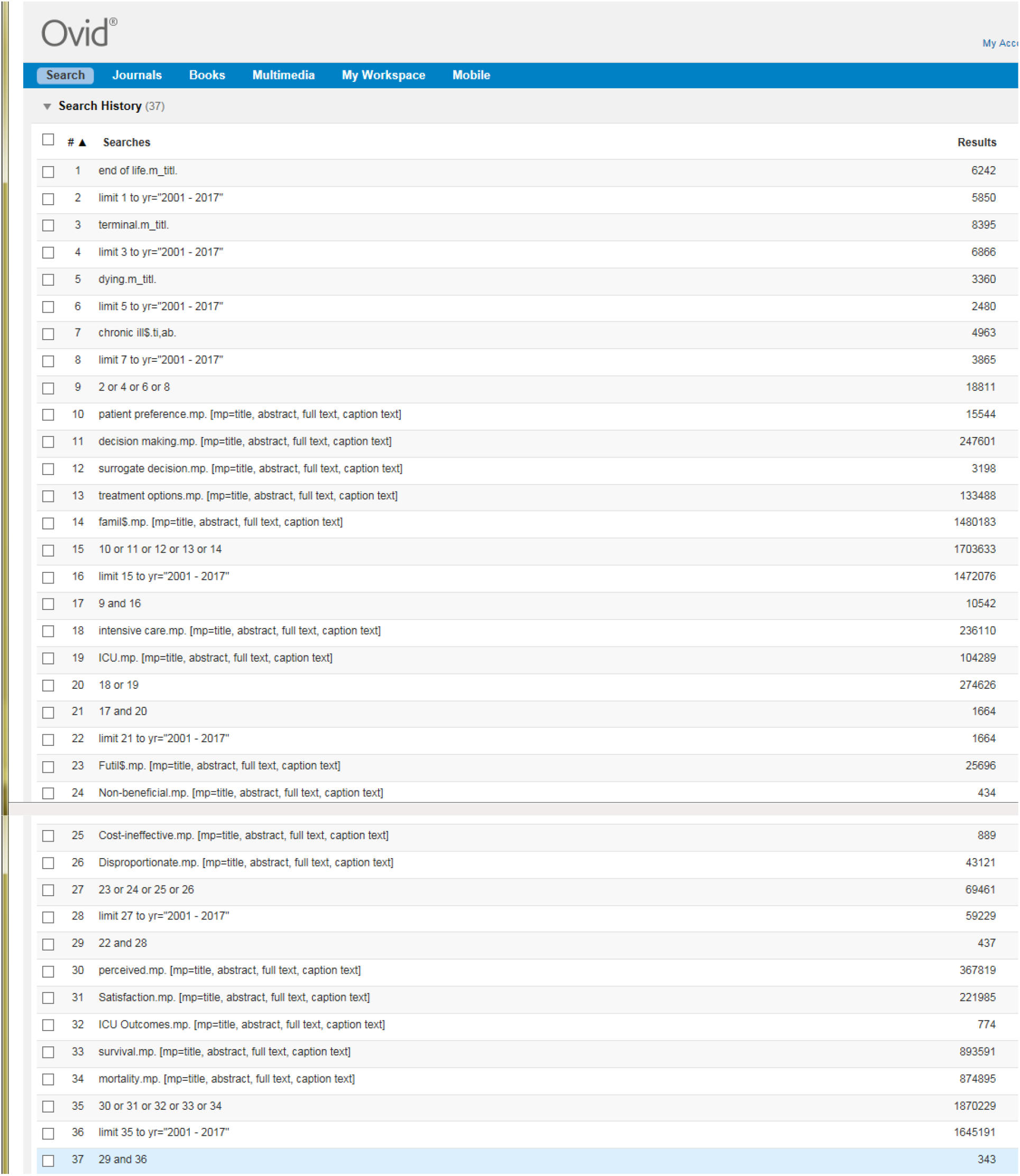

## PRISMA DIAGRAM

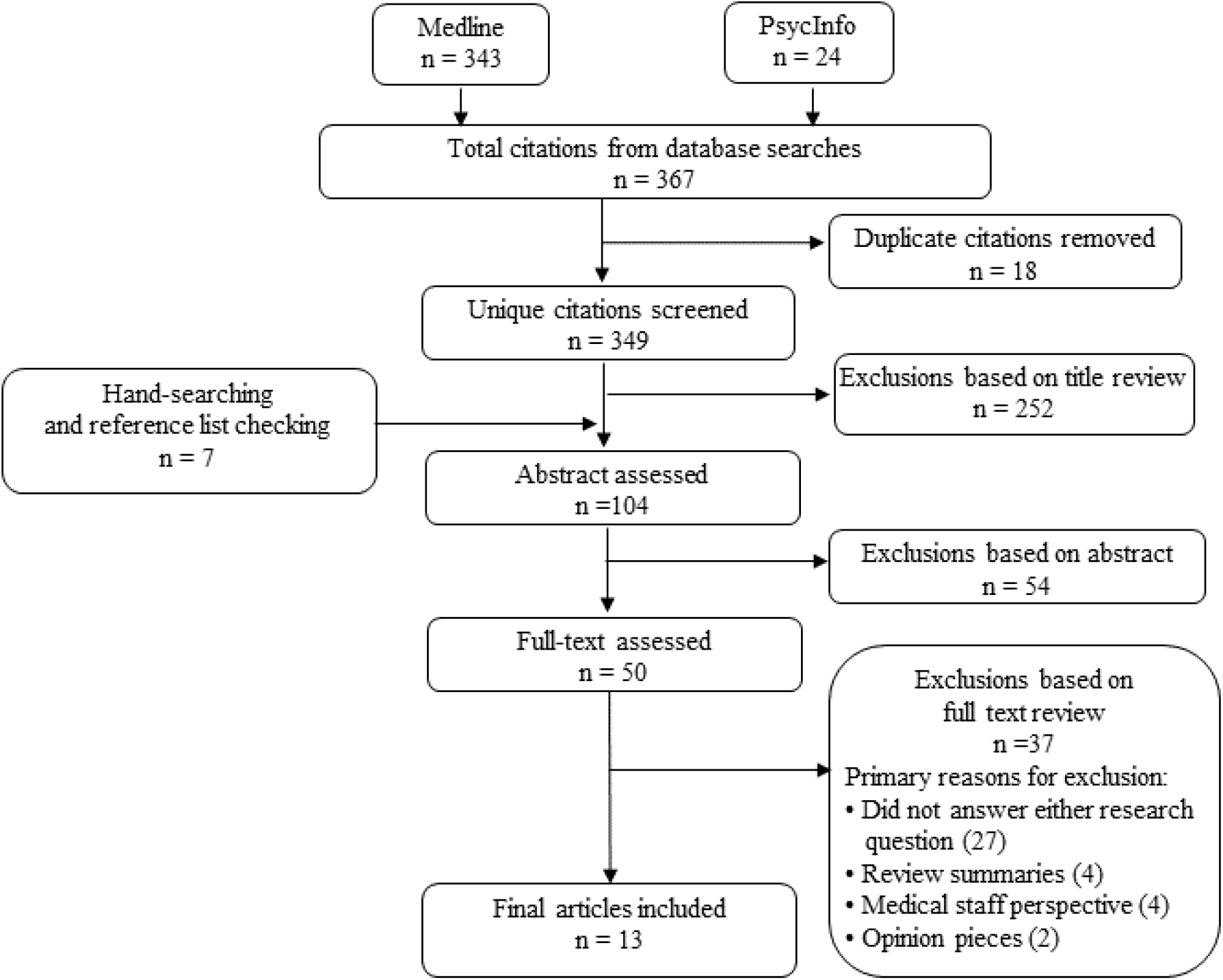

